# Study Pre-protocol for “BronchStart - The Impact of the COVID-19 Pandemic on the Timing, Age and Severity of Respiratory Syncytial Virus (RSV) Emergency Presentations; a Multi-Centre Prospective Observational Cohort Study”

**DOI:** 10.1101/2021.04.18.21255700

**Authors:** Thomas C. Williams, Mark D Lyttle, Steve Cunningham, Ian Sinha, Olivia Swann, Abigail Maxwell-Hodkinson, Damian Roland, on behalf of the Paediatric Emergency Research in the UK and Ireland (PERUKI)

## Abstract

**Background:** Bronchiolitis (most frequently caused by Respiratory Syncytial Virus; RSV) is a common winter disease predominantly affecting children under one year of age. It is a common reason for presentations to an Emergency Department (ED) and frequently results in hospital admission, contributing to paediatric units approaching or exceeding capacity each winter. During the SARS-CoV-2 pandemic, the circulation of RSV was dramatically reduced in the United Kingdom and Ireland. Evidence from the Southern Hemisphere and other European countries suggests that as social distancing restrictions for SARS-CoV-2 are relaxed, RSV infection returns, causing delayed or even summer epidemics, with different age distributions.

**Study question:** The ability to track, anticipate and respond to a surge in RSV cases is critical for planning acute care delivery. There is an urgent need to understand the onset of RSV spread at the earliest opportunity. This will influence service planning, to inform clinicians whether the population at risk is a wider age range than normal, and whether there are changes in disease severity. This information is also needed to inform decision on the timing of passive immunisation of children at higher risk of hospitalisation, intensive care admission or death with RSV infection, which is a public health priority.

**Methods and likely impact:** This multi-centre prospective observational cohort study will use a well-established research network (Paediatric Emergency Research in the UK and Ireland, PERUKI) to report in real time cases of RSV infection in children aged under two years, through the collection of essential, but non-identifying patient information. Forty centres will gather initial data on age, index of multiple deprivation quintile, clinical features on presentation, and co-morbidities. Each case will be followed up at 7 days to identify treatment, viral diagnosis and outcome. Information be released on a weekly basis and used to support clinical decision making.

## Introduction

Non-pharmaceutical interventions (NPIs) implemented to slow or eliminate the spread of SARS-CoV-2 have also had dramatic off-target consequences on transmission of other respiratory viruses, in particular Respiratory Syncytial Virus (RSV) in children. RSV infection can cause a clinical syndrome termed bronchiolitis, most commonly in infants under one year of age, and lower respiratory infection associated with wheeze in older children, typically under two years of age. In a typical year in England, RSV bronchiolitis causes significant morbidity, leading to an estimated 36,028 annual admissions for RSV, mainly in infants under 1 year of age;^1^ every year 0.1% of all infants under 1 are admitted to paediatric intensive care units (PICUs) with a diagnosis of bronchiolitis.^2^ In the years preceding 2020/21, RSV infections in the United Kingdom and Ireland saw consistent seasonal peaks in infection and hospitalisation, with cases generally starting to rise in September, peaking in November/December, and falling to summer levels by April.^1^ As such, when lockdown measures were implemented during the early spring of 2020, the RSV season was already in recession in the Northern hemisphere and no clear impact was seen on RSV disease burden. However, in the corresponding Southern Hemisphere winter of 2020, the NPIs led to dramatically reduced cases and admissions of paediatric RSV and influenza in Australia,^3^ New Zealand,^4^ Chile and South Africa.^5^ An associated reduced rate of admissions to Paediatric Intensive Care Unit (PICU) across South America, mainly driven by a reduction in respiratory diagnoses, was also observed.^6^

Hierarchical variance of NPIs over time have interrupted transmission of some, but not all, respiratory viruses.^7^ In the United Kingdom, test positivity for RSV in the Respiratory DataMart system has remained at less than 1% throughout the winter of 2020/21 (last data 11 April 2021).^8^ Conversely, rhinovirus infections continued throughout the Southern and Northern Hemisphere winter seasons.^8,9^ A typical annual spike in rhinovirus infections in children was observed with the return to schools in England in September 2020,^8^ and PICUs saw ongoing admissions for rhinovirus induced wheeze and bronchiolitis in the autumn/winter of 2020 despite lockdown measures [NHS Lothian data, unpublished].

Both observational and modelling studies suggest that RSV will arise out of season in the Northern hemisphere and may have a greater clinical impact.^10^ Four Australian states, which after an initial lockdown imposed few physical distancing measures with re-opened schools, have now experienced delayed annual RSV epidemic peaks. The delayed peak in Western Australia, where infections rates surged during local Spring-Summer (September-December), was associated with even higher case rates than normally seen during winter, despite no change in testing practices.^11^ Some European countries such as France have already seen evidence of a delayed RSV season, with all French regions now having reported bronchiolitis case rates at epidemic levels.^12^

From a clinical perspective, it is not possible to distinguish the viral aetiology of lower respiratory tract infection, with the exception of greater probability by season of infection. With the uncoupling of RSV and seasonality observed as an impact of NPIs, there is an urgent requirement to track RSV infection for season onset and to observe the impact on a larger unexposed at risk population. It is likely that when RSV returns to the United Kingdom and Ireland in 2021 it will not follow its usual predictable seasonal pattern, particularly if lockdown measures are relaxed rapidly as SARS-CoV-2 immunisation coverage increases. Reasons why RSV might display unusual disease dynamics include a lack of circulation in the general population leading to a reduction of transplacental antibodies and passive immunity for neonates, and the fact that children born after February 2020 will most likely not have been exposed to circulating RSV, thus reducing adaptive immune response to the virus when encountered. RSV infection is also associated with recurrent wheeze as a longer-term post infectious morbidity. A higher proportion of children affected by RSV may increase the subsequent healthcare burden of post infectious wheeze giving a much longer-term burden on health services.

The objective of this study is to monitor RSV disease in children under two years of age attending emergency departments across the UK and Ireland and examine the impact on timing, age and severity of clinical presentations as NPI restrictions are reduced throughout the UK and Ireland in 2021.

## Study Protocol

This protocol is structured in keeping with the principles of the STROBE statement (strobe-statement.org/).

### How the sample is to be selected

A national multi-centre prospective observational cohort study will be carried out through the PERUKI (Paediatric Emergency Research in the UK and Ireland) Network^13^ with forty departments having committed to participating in the study. All children meeting the inclusion criteria below will be eligible.

### Inclusion criteria

Children under two years of age presenting to participating emergency departments with clinical features of bronchiolitis (cough, tachypnoea or chest recession, and wheeze or crackles on chest auscultation)^14^ or a first episode of acute viral wheeze.

### Exclusion criteria

Children with previous episodes of wheeze responsive to bronchodilator, suggesting an underlying diagnosis of recurrent wheeze of early childhood.

### Data collection

We will collect data at two time points: baseline (date of presentation to a participating ED) and seven days later.

Clinicians identifying a case for inclusion will keep a local log of participants that contain patient identifiable characteristics cross referenced to a study number. The study number alone is used in all communication/data entry with the study team. An email after 7 days to the submitting clinician will prompt data entry at this point.

Anonymised data will be entered to a secure online database (REDCap data capture tool)^15,16^ (see below), by clinicians providing acute care within the ED.

### Variables to be measured

At baseline, data including patient demographics, presenting characteristics, acuity, investigations and treatments will be obtained (see Supplementary File 2), as will data on the number of siblings and travel. An external link will enable clinicians to enter a full postcode derived index of multiple deprivation score for database entry.

At 7 days data will include the child’s ultimate outcome (discharged or admitted), highest acuity dependency (the ward they were placed on if admitted: Normal, High Dependency or Intensive Care), whether care is ongoing, patient was discharged or died and (if obtained) what viruses were identified by PCR (see Supplementary File 3).

RSV status will be identified by either Nasopharyngeal Aspirate (NPA) (a) point of care testing (rapid viral testing where available) at baseline presentation to ED, or (b) by laboratory PCR testing, if either is performed as part of standard care.

### Sample size calculation

This is a convenience sample cohort study and as such a sample size is not calculated. However, the most recent bronchiolitis peak in France (epidemiological week 13, to 8 April 2021), involved 2,433 emergency department (ED) attendances and 953 hospital admissions in children under the age of 2 years, with a total of ∼16,000 ED attendances and ∼6,000 admissions since the start of 2021.^12^ The combined population of the United Kingdom and Ireland is 72 million, roughly equivalent to that of France (67 million). The PERUKI sites already enrolled in this study are estimated to provide care for 25 % of the total paediatric population, and if a similar trend is seen to France we would therefore expect to recruit 4,000 infants and children in the build up to the peak of an RSV epidemic. By site this would be a mean of ∼100 patients over 3 months, or approximately a patient a day per site. Previous PERUKI surveillance studies have recruited > 1,000 patients in short time scales (1-2 months).

### Study registration

The trial will be prospectively registered on ISCRTN Registry (https://www.isrctn.com/).

### Primary outcomes to be measured, as well as a list of secondary outcomes

#### Primary outcome

To report the timing, frequency and clinical severity of RSV infection in children under two years of age presenting to hospitals in the UK and Ireland.

#### Secondary outcomes

i. To report the excess morbidity of clinical presentations at ED with respiratory disease by inclusion criteria by month of presentation (compared to published literature/health data sources).
ii. To report the geographical timing and spread of RSV infection across the UK.
iii. To identify differences in severity of illness and patient population affected by RSV in relation to typical seasonal ranges in the published literature.
iv. To provide data to support and corroborate current RSV surveillance systems (i.e. Respiratory Datamart https://www.gov.uk/government/statistics/national-flu-and-covid-19-surveillance-reports) and the Emergency Department Syndromic Surveillance System (EDSSS). (https://www.gov.uk/government/publications/emergency-department-weekly-bulletins-for-2021).
v. To provide data to support decision making by government agencies (Joint Committee on Vaccination and Immunisation) in relation to the prescription of out of season Palivizumab for high risk children.
vi. To provide data to support estimates of potential longer-term impact of changes in severity of illness and patient populations on health (long term recurrent wheeze) and associated healthcare resources.

#### Data analysis and statistical plan

i. Demographics, regional frequencies, incidence estimates and logistic regression will be performed according to a Statistical Analysis Plan (SAP) in place prior to full data analysis. This will include:
ii. *Descriptive statistics*
  a. Weekly incidence (broken down nationally and regionally) calculated for population at risk for each Emergency Department.
  b. Median age, co-morbidities, index of multiple deprivation, presenting features of recruited patients, compared to previous population studies to identify if similar age, spectrum of co-morbidities and presenting clinical features to previous epidemics.
  c. Proportion of RSV vs non-RSV of those tested.
iii. *Identification of risk factors for severe disease*
  a. A number of metrics will be examined as proxy measures for severity including:
    i. Median saturations in air at presentation and frequency of attendance with SpO2 <94% or <92% in room air.
    ii. Median respiratory rate (with reference to age as per Paediatric Early Warning Score) at presentation.^17^
    iii. Blood gas composition (median H+ and pCO2 measurements) at presentation.
    iv. Proportion of children admitted to hospital and of those the proportion admitted to standard care / HDU / PICU.
    v. Proportion of children requiring respiratory support (supplementary oxygen, high flow oxygen, CPAP, BiPAP or invasive ventilation).
    vi. Length of stay.
  b. Multivariate logistic regression analyses using random effects model (recruiting site, availability of on-site PICU facility) to examine risk factors for hospitalisation, level of ventilatory support and PICU admission. Risk factors will be compared to predictors of severe disease published in population studies previously.
iv. *Spatio-temporal dynamics of the 2021 epidemic*
  a. Travel histories will be used to identify whether RSV is seeded into community in a number of separate introduction events, or spreads locally.

### Details of any ethical issues relating to the study (and of the ethical approval received)

This prospective observational surveillance study needs to be able to rapidly collate data that will enable other researchers and health services to make informed decisions about the extent of any RSV surge. For this reason, and the fact that the data being obtained is already obtained as part of the clinical record, we will not be asking for consent from families and carers. The clinician will only enter data regarding the outcomes of the child and no identifying information will be recorded. The patient’s study number will be linked to the local hospital number to allow outcome data to be retrieved. This information will be held by the recruiting site only in secure research lockers in each Emergency Department and only available to appropriately trained staff or research nurses.

None of the data recorded centrally by the study team will mean the patient is identifiable, and there are no specific details recorded that would make it possible to determine any individual child by proxy. This approach has been successfully adopted in other rapidly deployed studies the authors have been involved in. The security of the REDCap system we are using to collate data will add additional data protection safeguards.

The study will be submitted for Integrated Research Application System (IRAS) approval with University Hospitals of Leicester NHS Trust as the Study Sponsor, IRAS ID 297802.

### Data input, storage and management

Anonymised data will be entered using the validated online data entry software REDCap. (Research Electronic Data Capture tools) following the clinical report forms provided in the appendix (Supplementary Files 2 and 3). This software (REDCap) is hosted on the University Hospitals Bristol and Weston NHS Foundation Trust (UHBW) secure server, accessible on the Health and Social Care Network (HSCN) that is managed by NHS Digital. All research data reside within the hosting institution, and there will be a data transfer agreement made between University Hospitals of Leicester NHS Trust, Edinburgh University and University Hospitals Bristol and Weston NHS Foundation Trust in order to allow transfer of data on completion of data entry and cleaning. Processing of data will be undertaken on instruction of the Data Controller. The study Sponsor organisation (University Hospitals of Leicester NHS Trust) will be the Data Controller throughout, and University Hospitals Bristol and Weston NHS Foundation Trust will have Joint Controllership for the duration of data entry and cleaning. REDCap uses a granular security model so that users can only review the data they have been explicitly authorised to access. REDCap also provides a comprehensive log/audit feature that records all individual changes with a date/time stamp and a change owner. Data that are captured and stored will only be available via the information technology systems linked to the HSCN which is the current validated system used by NHS Trusts to share and store patient information.

### Plans for dissemination of the study outcome (including the associated data) once completed

Data will be presented in:

i. Data submissions to the regulatory authorities and local study teams, including an interactive online dashboard.
ii. Conference presentations.
iii. Peer reviewed scientific journals.
iv. Collaboration with educational websites with international social media reach (www.dontforgetthebubbles.com).
v. Engagement with the RSV patient network, part of the RESCEU Consortium (http://resc-eu.org/).

## Conclusions

There is an urgent need to better understand the effects of non-pharmaceutical interventions on the transmission and disease dynamics of Respiratory Syncytial Virus, a major cause of morbidity in infants in the United Kingdom. BronchStart is a multi-centre observational study embedded within the PERUKI network which will inform the immediate clinical response to a delayed RSV peak, improve our understanding of how RSV is seeded and spreads within the United Kingdom, and inform the implementation of future RSV immunisation campaigns.

## Data Availability

As this is a study protocol no data are yet available. Once available, anonymised, non-patient identifiable data will be stored on a RedCap server hosted by University of West of England, Bristol, United Kingdom (See Data input, storage and management).

## Ethics policies

The study protocol has been submitted to Integrated Research Application System; in view of the urgency needed to take this project forward, and the fact that no patient identifiable information will be collected, we believe it to be exempt from Research Ethics Committee (REC) Review. This is also in keeping with the Control of Patient Information Notice (Covid-19 – Notice under Regulation 3(4) of the Health Service Control of Patient Information Regulations 2002) last reviewed 3rd February 2021.

## Author contributions

Williams TC: Conceptualization, Methodology, Writing – Original Draft Preparation, Writing – Review & Editing

Lyttle MD: Conceptualization, Methodology, Project Administration, Software, Writing – Original Draft Preparation, Writing – Review & Editing

Cunningham S: Conceptualization, Methodology, Project Administration, Writing – Original Draft Preparation, Writing – Review & Editing

Sinha I: Conceptualization, Methodology, Writing – Original Draft Preparation, Writing – Review & Editing

Swann O: Conceptualization, Methodology, Writing – Original Draft Preparation, Writing – Review & Editing

Maxwell-Hodkinson A : Conceptualization, Methodology, Writing – Review & Editing

Roland D: Conceptualization, Methodology, Project Administration, Writing – Original Draft Preparation, Writing – Review & Editing

PERUKI Site Leads: Conceptualization, Writing – Review & Editing

## Acknowledgements

The authors would like to thank Mai Bacquedano for technical support in the launch of the REDCap survey tool and ongoing data management.

## Supplementary material

Supplementary File 1 lists the PERUKI investigators who will be coordinating local data entry.

Supplementary File 2 and 3 show the full questionnaires to be completed by clinicians/researchers, including the logic branching trees for questions asked.

