## Supplementary File 1 for "Study Pre-protocol for “BronchStart - The Impact of the COVID-19 Pandemic on the Timing, Age and Severity of Respiratory Syncytial Virus (RSV) Emergency Presentations; a Multi-Centre Prospective Observational Cohort Study”"

PERUKI Site Leads and Co-leads

| Site | Country | City | Site lead | Co-lead |
|---|---|---|---|---|
| Alder Hey Children's Hospital NHS Foundation Trust | England | Liverpool | Meriel Tolhurst-Cleaver | |
| Barking, Havering & Redbridge University Hospitals NHS Trust | England | Romford | Caroline Ponmani | |
| Birmingham Children's Hospital | England | Birmingham | Stuart Hartshorn | |
| Bolton NHS Foundation Trust | England | Bolton | Jessica Watson | |
| Bristol Royal Hospital for Children | England | Bristol | Roisin Begley | |
| Chelsea and Westminster NHS Foundation Trust | England | London | Sakura Hingley | |
| Countess of Chester NHS Foundation Trust | England | Chester | Steve Brearey | |
| Croydon University Hospital | England | Croydon | Darren Ranasinghe | |
| Evelina London Children’s Hospital | England | London | Sylvester Gomes | |
| Frimley Park Hospital | England | London | Patrick Aldridge | |
| Great North Children's Hospital, Newcastle Upon Tyne | England | Newcastle | Mark Anderson | |
| Hull Royal Infirmary | England | Hull | Elizabeth Herrieven | |
| Ipswich Hospital | England | Ipswich | David Hartin | |
| James Cook University Hospital | England | Middlesbrough | Kat Jerman | Arshid Murad |
| King's College Hospital, London | England | London | Rachael Mitchell | Fleur Cantle |
| Leicester Royal Infirmary | England | Leicester | Damian Roland | |
| Medway Hospital NHS Foundation Trust | England | Gillingham | Adebayo Da Costa | Alfred Sime |
| North Middlesex Hospital | England | London | Adam Lawton | |
| Northwick Park Hospital | England | Harrow | Bhavni Shah | Lauren Fraser |
| Nottingham University Hospitals NHS Trust | England | Nottingham | Ruth Wear | |
| Ormskirk & District General Hospital | England | Ormskirk | Sharryn Gardner | |
| Queen Elizabeth Hospital, Woolwich | England | London | Sharon Hall | |
| Royal Alexandra Children's Hospital | England | Brighton | Emily Walton | |
| Royal Derby Hospital | England | Derby | Gisela Robinson | Rachel Smith |
| Royal Wolverhampton NHS Trust | England | Wolverhampton | Lorna Bagshaw | |
| Salisbury NHS Foundation Trust | England | Salisbury | Seb Gray | |
| Sheffield Children's NHS Foundation Trust | England | Sheffield | Sally Gibbs | |

| | | | | |
|---|---|---|---|---|
| Shrewsbury & Telford NHS Trust | England | Shrewsbury | Lisa Kehler | |
| Southampton Children's Hospital | England | Southampton | Jane Bayreuther | |
| St George's Hospital, London | England | London | Heather Jarman | |
| St Mary's Hospital, Imperial College Healthcare NHS Trust | England | London | Neil Thompson | |
| The Royal London | England | London | Raine Astin-Chamberlain | |
| University Hospital Lewisham | England | London | Sophie Keers | |
| Watford General Hospital (West Herts NHS Trust) | England | Watford | Richard Burridge | |
| Bon Secours Hospital | Ireland | Cork | Ronan O'Sullivan | |
| Children's Health Ireland at Crumlin | Ireland | Dublin | Eleanor Ryan | |
| Children's Health Ireland at Tallaght | Ireland | Dublin | Sheena Durnin | Stanley Koe |
| Royal Belfast Hospital for Sick Children | N. Ireland | Belfast | Julie-Ann Maney | |
| Royal Hospital for Children & Young People, Edinburgh | Scotland | Edinburgh | Jen Browning | |
