## Supplementary File 2 for "Study Pre-protocol for “BronchStart - The Impact of the COVID-19 Pandemic on the Timing, Age and Severity of Respiratory Syncytial Virus (RSV) Emergency Presentations; a Multi-Centre Prospective Observational Cohort Study”"

### Bronchiolitis Surveillance - ED attendance details

The BronchSTART study is capturing information on patients with a clinical diagnosis of bronchiolitis or first episode of wheeze. Contrary to normal practice for bronchiolitis studies in the UK & Ireland, our upper age limit is the second birthday, as due to reduced exposure, older children may be prone to RSV infection.

This form captures information from the Emergency Department (or similar unit) attendance. If that isn't finished by the time you finish your shift, you can return to this when you're next on duty - BUT PLEASE REMEMBER TO SAVE THE RETURN CODE. Your local BronchSTART study lead will be able to tell you how to generate a study ID for your patient.

Please enter your work email address, so that we can send you the short follow up form seven days after this attendance. Please be assured, all details, including your email address, are stored on NHS servers in UHBW NHS FT, and the information you provide will not be used for any purpose other than the BronchSTART study.

Thanks a million for submitting this form – with all of us working together, we will be able to spot early rises in RSV across the country, allowing us to adjust clinical services, and inform future vaccine programmes and research studies. If you run into problems at any time, please email the study team at

#### Eligibility check, and submitter details

| | |
|---|---|
| Is the child aged under two years? | ◯ Yes<br>◯ No |
| Does this child have a clinical diagnosis of bronchiolitis, acute lower respiratory tract infection or FIRST episode of wheeze? | ◯ Yes<br>◯ No |
| Has this child previously been enrolled in this surveillance study?<br>(please check your local study log if unsure) | ◯ Yes, within the last month<br>◯ Yes, but more than one month ago<br>◯ No |

This child is eligible - please enter their participation number in the following field

This child is ineligible - please do not proceed any further

Please see your local study log for the next number to be allocated - this should consist of your three letter site code, and a three digit number

As it is more than one month since this child was previously enrolled, please enrol them again, with a NEW STUDY NUMBER

Please see your local study log for the next number to be allocated - this should consist of your three letter site code, and a three digit number

| | |
|---|---|
| Please enter the study code for this patient here<br>(Please see your local study log for the next number to be allocated - this should consist of your three letter site code, and a three digit number) | ______________________ |
| What is YOUR first name? | ______________________ |
| Please enter the email you use for patient-related work purposes here | ______________________ |

#### Attendance & patient details

Date of current ED attendance ______________________

How old is the child in months? ______________________

How old is the child in weeks? ______________________

Sex of child
- ◯ Male
- ◯ Female

Does the child have any co-morbidities?
- ☐ Prematurity (< 37 weeks)
- ☐ Chronic Lung Disease of prematurity
- ☐ Congenital Cardiac Disease
- ☐ Neuromuscular Disease
- ☐ Other
- ☐ None

Gestational age at birth
(round down to completed weeks - if not known, document NK)
- ◯ 22
- ◯ 23
- ◯ 24
- ◯ 25
- ◯ 26
- ◯ 27
- ◯ 28
- ◯ 29
- ◯ 30
- ◯ 31
- ◯ 32
- ◯ 33
- ◯ 34
- ◯ 35
- ◯ 36
- ◯ Not known

Has the child had palivizumab within the last month?
- ◯ Yes
- ◯ No
- ◯ Unknown

What other comorbidities does the child have? ______________________

How many siblings does this child have?
(if not known, please document NK)
- ◯ 1
- ◯ 2
- ◯ 3
- ◯ 4
- ◯ 5
- ◯ 6
- ◯ 7
- ◯ 8
- ◯ 9
- ◯ 10
- ◯ 11
- ◯ 12
- ◯ Not known

| | |
|---|---|
| Has the child or anyone in the household travelled further than their "local" environment in the last six weeks?<br><br>(for example, travelling into another county/region - provided they don't live right on the border) | ◯ Yes<br>◯ No<br>◯ Not known |
| To which geographical region(s) did they travel? | ☐ UK<br>☐ Europe<br>☐ Other |
| Please name the destination here | __________________________________ |

#### Investigations

What was the first recorded oxygen saturation level?

__________________________________

Was this in:

- ◯ Air
- ◯ Supplemental oxygen

What was the first recorded respiratory rate?

__________________________________

Were any of the following investigations performed in the Emergency Department?

- ☐ Point of care virus testing
- ☐ Blood Gas
- ☐ None

Was the child

- ☐ RSV positive
- ☐ Other virus positive
- ☐ Virus negative

What other virus(es) was the child positive for?

__________________________________

What type of blood gas sample was this?

- ◯ Capillary/Arterial
- ◯ Venous
- ◯ Not known

Acid-base unit

- ◯ pH
- ◯ H+ (mmHg)

What was the pH on the blood gas?

__________________________________

What was the H+ mmHg result on the blood gas?

__________________________________

PaCO2 unit

- ◯ kPa
- ◯ mmHg

What was the PaCO2 in kPa?

__________________________________

What was the PaCO2 in mmHg?

__________________________________

#### Treatment & disposition

| | |
|---|---|
| Did the child require any of the following while in the Emergency Department? | ☐ Suction<br>☐ Feed/Fluid support<br>☐ Oxygen/respiratory support<br>☐ None of the above |
| Which mode(s) of feeding/fluid support were required in the Emergency Department? | ☐ Nasogastric fluids<br>☐ Intravenous fluids |
| Which mode(s) of oxygen/respiratory support were required in the Emergency Department? | ☐ Oxygen - Low flow (face mask/nasal cannulae/headbox)<br>☐ Oxygen via High Flow Humidifying Device<br>☐ CPAP or BIPAP<br>☐ Invasive mechanical ventilation |
| On completion of the Emergency Department episode, the outcome was: | ○ Discharged home<br>○ Admitted to a Short Stay or Observation Unit<br>○ Admitted to an Inpatient Ward<br>○ Admitted to a HDU<br>○ Admitted to a PICU<br>○ Died in Emergency Department |
