## Supplementary File 3 for "Study Pre-protocol for “BronchStart - The Impact of the COVID-19 Pandemic on the Timing, Age and Severity of Respiratory Syncytial Virus (RSV) Emergency Presentations; a Multi-Centre Prospective Observational Cohort Study”"

### Bronchiolitis surveillance follow up

Dear [name], thanks very much for completing this seven-day follow up form.

Please note: while we want information relating to seven days after the initial attendance, you do not need to complete this form on that exact day.

If you need to leave this form and come back to it later, you can do so - but please remember to save the return code, and enter your email address when prompted

**This is the seven-day follow-up form for study participant [study_id], who initially attended on [attend_dat]**

**If you are trying to do anything other than enter follow-up information for this patient, please leave this form now**

What date is seven days after the initial attendance? __________________

Days between surveys __________________

The date you have entered is not seven days after the initial attendance - please recheck this

#### Follow up details

| | |
|---|---|
| Please enter the IMD score for the patient's postcode<br><br>(click here to open the IMD tool in a separate window) | ◯ 1<br>◯ 2<br>◯ 3<br>◯ 4<br>◯ 5<br>◯ 6<br>◯ 7<br>◯ 8<br>◯ 9<br>◯ 10<br>◯ Not known |
| At midday on [fu_dat], was the patient: | ◯ At home<br>◯ In an inpatient ward<br>◯ In an HDU<br>◯ In a PICU<br>◯ Deceased |
| On [attend_dat], you stated the patient had been [disposition] - what happened since then? (select all that apply) | ☐ Returned to ED, but discharged home from ED again<br>☐ Returned to ED, and admitted to hospital<br>☐ Remained discharged with no further ED contact |
| On [attend_dat], you stated the patient had been [disposition] - what happened since then?<br><br>(select all that apply) | ☐ Remained an inpatient throughout<br>☐ Subsequently discharged, returned to ED and readmitted<br>☐ Subsequently discharged, returned to ED and discharged home<br>☐ Subsequently discharged, no further ED contact |
| What was the total number of attendances during the seven day period<br><br>(including the index attendance) | ◯ 1<br>◯ 2<br>◯ 3<br>◯ 4<br>◯ 5<br>◯ 6<br>◯ Not known |
| Please select all areas to which the child was admitted during their hospital stay(s) | ☐ Observation Unit<br>☐ Inpatient Ward<br>☐ High Dependency Unit<br>☐ Intensive Care Unit<br>☐ Other |
| Did the child have an RT-PCR test during this illness? | ◯ Yes<br>◯ No |
| What was the result of the RT-PCR test? (select all that apply) | ☐ RSV positive<br>☐ Other virus positive<br>☐ Virus negative |
| What other virus(es) was the child positive for? | ☐ Adenovirus<br>☐ Human Metapneumovirus<br>☐ Influenza<br>☐ Parainfluenza<br>☐ Rhinovirus<br>☐ SARS-CoV-2<br>☐ Other |

| | |
|---|---|
| Please list any other virus(es) here | __________________________________ |
| Which of the following treatments were required during the hospital admission(s)?<br>(select all that apply) | ☐ Nasogastric fluids<br>☐ Intravenous fluids<br>☐ Oxygen - Low flow (fask mask/nasal cannulae/Headbox)<br>☐ Oxygen via High Flow Humidifying Device<br>☐ CPAP or BIPAP<br>☐ Invasive mechanical ventilation<br>☐ Pharmacologic treatment(s)<br>☐ None of the above |
| Which of the following pharmacologic treatments were given during the hospital admission(s) | ☐ Salbutamol<br>☐ Other bronchodilator<br>☐ Prednisolone<br>☐ Antibiotic<br>☐ Other |
| Which other bronchodilator treatment was given? | __________________________________ |
| Which other pharmacologic treatments were administered? | __________________________________ |
